# Video-evoked neuromarkers of visual function in age-related macular degeneration

**DOI:** 10.1101/2025.02.13.25322213

**Authors:** Angela I. Renton, David J. Klein, Jesse A. Livezey, Dan Nemrodov, Stephanie Wolfer, Adam Hanina, Dimitri Van De Ville

## Abstract

Neural markers of visual function in age-related macular degeneration (AMD) allow clinicians and researchers to directly evaluate the functional changes in visual processing which occur as a result of the progressive loss of afferent input from the macula. Unfortunately, few protocols exist that elicit such neural markers, and most of these are poorly adapted to AMD. Here, we propose a novel method of embedding frequency tags into full colour and motion videos by periodically manipulating the contrast of visual information of different spatial frequencies at different temporal frequencies. These videos elicit steady-state visual evoked potentials (SSVEPS) in viewers which, when measured using electrophysiological neuroimaging methods, independently represent the responses of populations of neurons tuned to the tagged spatial frequencies. We used electroencephalography (EEG) to record the SSVEPs of 15 AMD patients and 16 age-matched healthy controls watching a 6-minute series of natural scene videos filtered with this spatial frequency tagging method. Compared with healthy controls, AMD patients showed a lower SSVEP response to high spatial frequency information, and a stronger response to the low spatial frequency information in the video set. The ratio of the SSVEP response to lower relative to higher spatial frequency information was strongly predictive of both visual acuity and contrast sensitivity, and the topographic distributions of these responses suggested retinotopic reorganisation of the neural response to spatial frequency information.

## INTRODUCTION

Age related macular degeneration (AMD) is the most common cause of vision loss in older adults, affecting 8.7% of people aged 45-85 years globally (Stahl, 2020). This amounted to an estimated 196 million people suffering from AMD in 2020, with a projected increase to 288 million people by 2040 (Wong et al., 2014). This chronic disease affects the macular region of the retina and is characterized by progressive loss of central vision (Lim et al., 2012; Rattner and Nathans, 2006). AMD can severely impinge on patient’s quality of life and independence by interfering with visual discrimination tasks such as reading (Varadaraj et al., 2018), face and emotion recognition (Boucart et al., 2008; Logan et al., 2022), object and scene recognition (Boucart et al., 2013; Thibaut et al., 2018), interaction with electronic devices (Taylor et al., 2016), and driving (Rovner and Casten, 2002; Wood et al., 2018). Clinicians and researchers who work to understand and treat AMD require accurate and comprehensive measures of AMD patients’ changing visual function. Central vision loss is known to result in structural and functional changes in the brain’s visual system, yet standard visual function tests rarely directly measure these changes (Baker et al., 2005; Broadhead et al., 2020; Chandramohan et al., 2016; Cheung and Legge, 2005; Ramanoël et al., 2018). In turn, researchers still have a limited understanding of what adaptive versus maladaptive changes in neural activity look like in the face of central vision loss (Rosa et al., 2013). To address this gap, we developed and tested a novel neuroimaging-based visual function test for AMD which harnesses visual evoked neural responses to full-colour natural scene videos.

The macular region of the retina is characterised by a high density of both light sensitive photoreceptor cells and the retinal ganglion cells they innervate, with the density of these photosensitive cells decreasing systematically toward the peripheral visual field. The macula thus senses changes in light and colour on a fine scale, allowing the brain to resolve visual details such as the shapes of letters on a page (Ambati and Fowler, 2012). Spatial frequency, the rate at which information changes over space, is therefore differentially detectable across the visual field, with the highest spatial frequencies detectable in central vision (Drasdo et al., 2007; Hadjikhani and Tootell, 2000; Ramanoël et al., 2018). In turn, spatial frequency sensitivity in the brain’s visual system is retinotopically mapped, such that neurons sensitive to higher spatial frequencies are more populous in regions of the cortex responsive to the central visual field, and neurons sensitive to lower spatial frequencies are more populous in regions of the cortex mapped to the periphery of the visual field (Kauffmann et al., 2014). The detection of contrasted edges at distinct spatial frequencies is one of the first steps in hierarchical visual processing; with this information feeding forward through the visual system to allow for more complex perceptual processing of visual information (Tovée, 1996). As such, the deafferentation of visual cortical tissue mapped to the macular region of the visual field should be expected to have a profound effect on neural activity in the brain’s visual cortex.

There is substantial evidence suggesting that AMD patients undergo both structural and functional changes in vision. Several studies have shown that long-term AMD patients present with atrophy within cortical areas retinotopically mapped to central vision (Boucard et al., 2009; Prins et al., 2016) and cortical thickening in regions mapped to more peripheral regions of the visual field (Burge et al., 2016). The cortical thickening is suggestive of compensatory changes as visual perceptual processing adjusts to the loss of central visual input. Indeed, many functional studies in both animals and humans support this notion; finding evidence of retinotopic reorganisation following central vision loss (see Cheung and Legge, 2005 for a review). These changes can be seen even in short term adaptation to scotopic vision, such that neurons whose receptive fields overlap with a central scotoma shift their receptive fields to encompass a larger area centred more peripherally in the visual field (Barton and Brewer, 2015). This adaptive reorganisation has been shown to be more dramatic in long-term AMD patients, with visual cortical areas which map to central vision in healthy adults found to respond to peripheral visual stimulation in AMD patients (Baker et al., 2008, 2005; Dilks et al., 2014; Liu et al., 2010; Plank et al., 2021; Schumacher et al., 2008). Further evidence for functional reorganisation comes from the tendency of AMD patients to develop a preferred retinal locus (PRL); i.e., a location in the peripheral retina adopted as pseudo-fovea by the oculomotor system, setting a reliable and automatic new location for fixation (Crossland et al., 2011; Maniglia et al., 2023, 2020; Rees et al., 2005). Psychophysical testing has revealed that visual perceptual function in the visual field around the PRL mimics that of the visual field around the fovea in healthy vision (Chen et al., 2019; Chung, 2014). Together, these results strongly suggest that the visual system undergoes functional reorganization to accommodate the loss of central visual information from the retina.

The functional changes in neural activity found for AMD patients can be either adaptive or maladaptive, either allowing patients to optimise their remaining visual function or engendering ancillary visual perceptual pathologies. As an example of an adaptive change, Shanidze and Verghese, 2019 found that motion discrimination is well preserved in AMD patients. This is likely because motion sensitive areas of the visual cortex are typically innervated by the magnocellular pathway, which largely contains information from peripheral vision (Hadjikhani and Tootell, 2000). Crucially, the authors found a positive correlation between motion discrimination performance and the time since AMD diagnosis, suggesting that patients can adapt to improve their preserved visual function as their ability to discriminate central visual information wanes.

By contrast, up to 40% of ocular pathology patients develop a condition called Charles Bonnet syndrome as a result of the loss of part of their visual field (Teunisse et al., 1996). Charles Bonnet patients, who have no comorbid psychiatric conditions, experience vivid long-term hallucinations ranging from simple geometric shapes, patterns, and flashing lights to complex hallucinations of animals, faces and even entire scenes (ffytche, 2009; Santhouse et al., 2000; Singh et al., 2014). Painter et al., 2018 found compelling evidence to support a long-held hypothesis that Charles Bonnet syndrome in AMD can be attributed to cortical hyperexcitability, as Charles Bonnet AMD patients display strikingly elevated visual cortical responses to peripheral visual field stimulation compared with control AMD patients. These examples highlight the importance of visually evoked neural activity in forming a full picture of any AMD patient’s visual function.

To date, several tests of visual function have been developed that directly measure visually evoked neural activity. For example, steady-state visual evoked potential (SSVEP) measures of visual acuity use electroencephalography (EEG) to measure evoked neural responses to a sweep of simple pattern reversing grating stimuli across a range of spatial frequencies. Sweep protocols aim to harness these SSVEPs at the grating flicker-frequency to measure the threshold highest granularity of visual information that a patients’ visual cortex can resolve (Hamilton et al., 2021a). If a patient has lost the ability to resolve a grating of a particular high spatial frequency, the pattern-reversal flicker appears as a uniform grey surface and will not evoke an SSVEP. Unfortunately, while visual acuity scores derived with this SSVEP thresholding approach align well with objective behavioural measurements in general, this correlation is less strong for AMD patients (Hamilton et al., 2021b). Neuroimaging-based protocols that specifically aim to measure visual field losses rely on similar principles to sweep SSVEP protocols, typically presenting a dartboard-like checkerboard composed of concentric rings of black and white gratings. These methods measure neural response to pattern reversals in specific regions of the checkerboard stimulus (Bach, 2006; de Santiago et al., 2019; Horn et al., 2016; Liu et al., 2021). These techniques require patients to maintain fixation at the centre of the display, to ensures that patients’ visual fields line up correctly with the checkerboard stimulus. Unfortunately, this poses a significant barrier for AMD patients who struggle to maintain central fixation due to their central vision loss. Thus, current neuroimaging-based measures of visual function are relatively poorly adapted to AMD.

Currently available neuroimaging measures of visual function share two additional limitations in their applicability to AMD patients. The first is that they rely on detecting a threshold spatial frequency or location at which neural responses are no longer detectable. However, many visual impairments will lead to reduced, heightened or shifted (i.e., in time or location in the brain) neural responses rather than a simple absence of response. These more subtle shifts in neural activity are not measured by the standard application of currently available tests. The second limitation is that these protocols rely on simple monochrome visual stimuli, such as gratings and checkerboards. These high contrast stimuli optimally stimulate early visual cortical activity and maximise the signal-to-noise ratio or the measured neural response. However, these stimuli also have low ecological validity as test stimuli, and have been shown to be less effective than more complex visual stimuli in stimulating neural activity through purely extra-foveal stimulation(Nemrodov et al., 2024). Natural visual scenes contain colour and motion information and have a complex hierarchical mathematical structure; i.e., spatial frequencies are grouped across time and space following scale-invariant fractal geometry, and exhibit a 1/*f* power law (Zetzsche, 2005). The human brain is optimized to this structure. Further, due to both the anatomical structure of the eye and the functional organisation of the visual system, there are retinotopic spatial and temporal patterns to colour, motion, and spatial frequency sensitivity which cannot be detected using monochrome patterns. For example, lower spatial frequency information, largely sensed in peripheral vision, is conveyed to the visual cortex through a faster cellular pathway than the higher spatial frequency information detected in central vision (Johnson and Johnson, 2014; Neri, 2014; Roberts et al., 2022). Low spatial frequency information, processed earlier, is used to extract scene context which is fed back down the visual hierarchy to allow predictive guidance in the processing of high spatial frequency information (Kauffmann et al., 2014). This fast and automatic feedforward/feedback process strongly impacts neural responses to visual information of distinct spatial frequencies but cannot be measured using simple monochrome pattern stimuli.

Here, we developed and benchmarked a novel neuroimaging-based visual function test which aims to address limitations in past neuroimaging measures of visual function in AMD. We propose a novel method to embed frequency tags within full-colour and motion natural scene videos, such that the contrast of information at different spatial frequencies is periodically modulated at different temporal frequencies to elicit SSVEPs. Next, we propose a test of visual function in AMD which measures the changing sensitivity to spatial frequency information in these videos. This test relies on the retinotopic organisation of spatial frequency sensitivity and would thus allow for the identification of central visual field deficits while participants freely and naturally shift their gaze across the display. We hypothesise that the novel video spatial frequency tagging method will evoke SSVEPs, allowing for concurrent but independent measurement of the population responses of neurons tuned to each of the tagged spatial frequencies. Further, we hypothesise that compared with healthy, age-matched controls, AMD patients should present a reduced SSVEP response to higher spatial frequency information due to the loss visual sensitivity in central vision, but a larger response to lower spatial frequency information due to adaptive changes in visual perceptual processing. To assess these hypotheses, we created a stimulus set in which we tagged two spatial frequency ranges: a higher range which should only be resolvable given the density of retinal ganglion cells within paracentral vision (< 8°/visual angle within visual field, > 3.2 c/d), and a lower range to which cortical regions retinotopically mapped to peripheral vision should be most sensitive (<3.2 c/d, Metha and Lennie, 2001). In total 15 patients diagnosed with bilateral dry AMD and 16 age-matched healthy controls viewed a 6-minute series of these spatial frequency tagged videos while we recorded EEG data to allow for the measurement of SSVEP responses. In analysing these data, we found that the novel video tagging method elicited SSVEP responses at the tagged frequencies. In turn, we showed that these SSVEP responses were sensitive to the changes in visual processing associated with AMD. We therefore propose that video-evoked SSVEPs to spatial frequency information in natural scenes can be used as a neural marker of visual function in AMD.

## METHODS

### Participants

N = 15 patients diagnosed with bilateral dry AMD (13 females, age M = 74.75 years, SD = 6.81) and 16 healthy older adults (14 females, age M = 61.3 years, SD = 3.3) volunteered to participate in the experiment after providing informed consent. Healthy controls were recruited through Institutional Review Board (IRB)-approved recruitment materials including flyers, online advertising on social media platforms, and business cards. AMD patients were referred and pre-screened by an ophthalmologist to the research team who completed the screening and enrolment process. AMD patients were included based on their age (at least 50 years old), a confirmed diagnosis of bilateral Dry AMD by their ophthalmologist, visual acuity in both eyes between 20/30 and 20/100, Mini-Mental State Exam (MMSE) score of at least 25, and ability to provide written informed consent. Geographic atrophy (GA) was present in many, but not all of the AMD patients (see **Figure 1**). Healthy controls were included based on age (at least 50 years old), best corrected visual acuity better than 20/25, and no history of any eye or optic nerve conditions. Participants were screened for psychiatric neurological conditions that could affect their vision or cognitive abilities, photosensitivity to flickering images and lights and history of epilepsy or seizures, no recent eye exam within the last 2 years, evidence of wet AMD or active neovascular leakage, any implanted electronic devices, history of cardiac problems, and any active infection or inflammation in the eyes. AMD participants were paid $88/hr and healthy controls were paid $38/hr for their participation. Both AMD patients and controls performed this experiment as part of a larger battery of tests not presented here. The study was approved by the Biomedical Research Alliance of New York (BRANY) Ethics Committee, and the experiment was performed according to the relevant guidelines and regulations.

**Figure 1.**
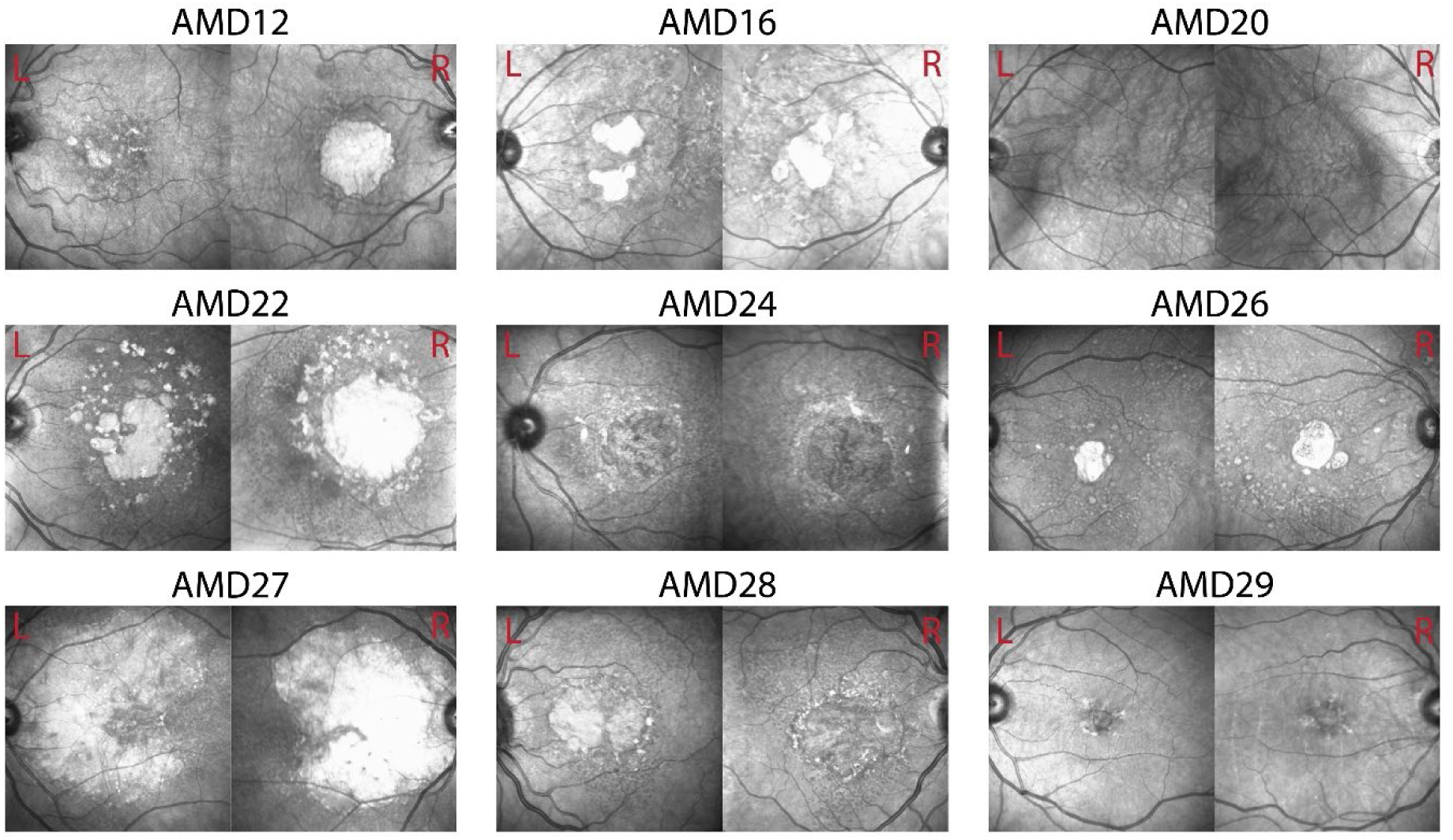
Optical coherence tomography (OCT) images of left and right eyes in example patients. Visual function scores for these patients can be found in Table 2.

### Experiment Design

All participants were instructed to freely view a 6-minute series of frequency-tagged natural-scene videos while we recorded EEG data. Participants were not instructed to fixate at any position on the display. The video set consisted of 6 unique, 7.5 second videos (see **Figure 2a** for a sample frame from each video or visit https://osf.io/rp4q5/ to view videos). For each of these videos, we frequency-tagged relatively higher and relatively lower spatial frequency information (>3.2 cyc/deg, < 3.2 cyc/deg), manipulating the contrast of information within these spatial frequency ranges at different rates (7Hz, 9Hz) to induce SSVEPs. We choose 3.2 cyc/deg as the threshold spatial frequency to which reginal ganglion cells retinotopically mapped to regions of the visual field outside of paracentral vision (> 8°/visual angle) should no longer be sensitive. Temporal frequency and spatial frequency (SF) were counterbalanced, such that each unique video was presented under two conditions (Condition 1: high SF - 7 Hz, low SF - 9 Hz | Cond 2: high SF: 9 Hz, low SF – 7Hz). The 6-minute series was therefore composed of each of the 6 unique videos, presented under each of the 2 tagging conditions, repeated 4 times per condition for a total 48 video presentations. Videos were presented in random order with no breaks between videos.

**Figure 2.**
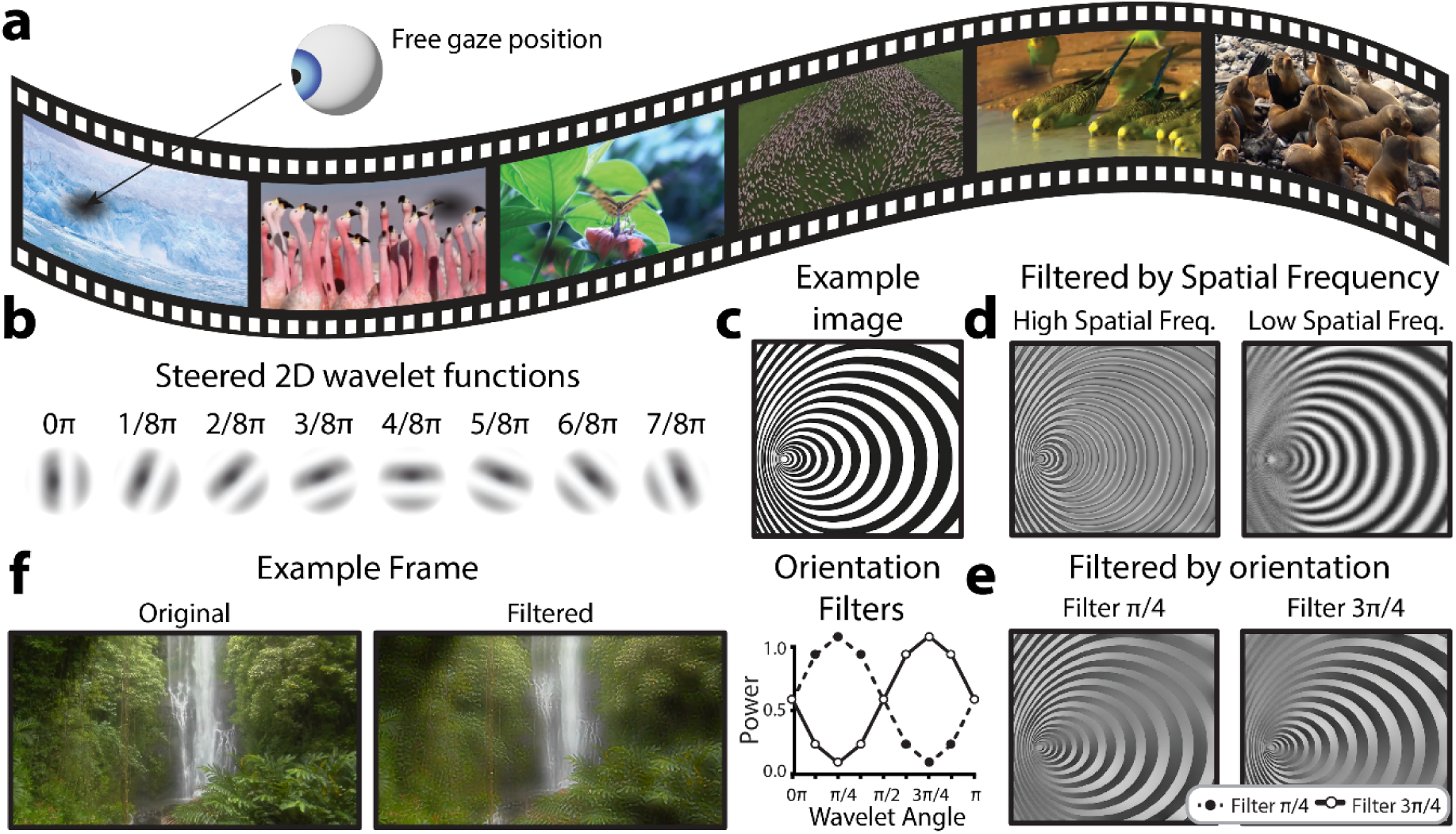
Overview of visual stimulus design. a) Sample frames of each of the 6, 7.5 second natural scene video clips used in the experiment. Participants were free to shift their gaze at will. b) Illustrative steered 2D wavelet functions used for signal embedding. Each frame was decomposed into 5 spatial frequency bands at 8 different orientations to allow the contrast of these visual features to be independently manipulated. c) By manipulating the power of different spatial frequencies and orientations in an image’s steerable wavelet pyramid, it is possible to reconstruct the image with altered contrast in specific orientation and spatial frequency bands. This illustration shows the effect of applying this approach to filter out d) relatively higher or lower spatial frequencies at all orientations, or e) each of the two oblique orientations at all spatial frequencies. f) An example video frame before and after filtering to embed the flicker signal. This frame highlights low spatial frequency information.

### Frequency tag embedding

The novel spatial frequency video tagging method presented here relies on embedding temporal frequency tags into the spatial frequency structure of natural scene videos. To achieve this, we applied a steerable 2D wavelet pyramid method to alter the power with which different spatial frequencies and orientations were represented in each frame (Karasaridis and Simoncelli, 1996; Simoncelli et al., 1992; Simoncelli and Freeman, 1995). This method was used instead of a conventional 2D Fast-Fourier Transform (FFT), because it allows for segmentation across both orientation and spatial frequency; thus, creating the possibility to flexibly choose how different orientations are tagged. Indeed, this method could also be applied purely to tag oriented information, regardless of spatial frequency. Steerable wavelet transforms involve convolving an input signal with a “mother wavelet” function, which is scaled (stretched wider and narrower) to capture the input signal at different spatial frequencies, and rotated to respond to different orientations at a given spatial frequency (Van De Ville and Unser, 2008). Here, we used a Simoncelli isotropic wavelet function as the mother wavelet (**Figure 2b**) to build the steerable wavelet pyramid, which provides good estimates for early visual processing (Portilla and Simoncelli, 2000). One of the computational benefits of this method is that the orientation-spatial frequency space representation is perfectly invertible, and thus it is possible to adjust the power with which different spatial frequencies and orientations are represented across the image and then reconstruct meaningful images incorporating these changes. For example, consider the image in **Figure 2c**, which varies in both spatial frequency and orientation over space, for illustrative purposes. Using the 2-D steerable wavelet pyramid, it is possible to reconstruct the image after suppressing the power within specific spatial frequency (**Figure 2d**) or orientation (**Figure 2e**) channels. When the image is filtered for high spatial frequencies (**Figure 2d**, left), high spatial frequency regions and edges throughout the image are emphasized and smooth, low frequency areas are filtered (averaged into grey). In contrast, when the image is filtered for low spatial frequencies (**Figure 2d**, right), high spatial frequency regions turn into grey and only the larger spatial structures are preserved. Similarly, areas with oriented edges are differentially preserved or filtered depending on the orientation of the filter (**Figure 2e**). The steerable wavelet decomposition for this project was applied using the Steerable Wavelet Transform Toolbox in MATLAB (Püspöki and Unser, 2015; Unser and Chenouard, 2013). We recommend the Plenoptic toolbox for implementation in Python, which we have used successfully for the same purpose in other projects (Balzani et al., 2024).

To apply this method, we converted each full-colour video frame to hue, saturation, value (HSV) colour space and extracted the value (luminance) channel (see example single frame in **Figure 2f**). We parsed this luminance information to a 2D steerable wavelet pyramid to extract the power within 5 spatial frequency bands for 8 orientations (0π – 7π/8, **Figure 2b**). This allowed us to separate out spatial frequency information larger and smaller than the threshold (3.2 cyc/degree). For each of these spatial frequency ranges, we sought to specifically tag information at oblique (rather than cardinal orientations). In general, cardinally oriented information is more prevalent in natural scenes, and in turn the visual system is more sensitive to visual information at cardinal orientations (Tovée, 1996). Thus, applying the embedded tag to obliquely orientated information allowed us to probe neural responses to distinct spatial frequencies while minimising the semantic distortion to the natural scene videos. This method could also be applied to tag cardinal orientations to elicit a stronger response. To achieve the tagging of oblique information, we applied a filter to scale the power across the 8 recovered orientations (for the relevant spatial frequency band) according to a phase of the embedded frequency tag (7 or 9 Hz sin wave). The power filter F was computed as follows:

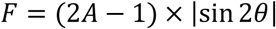

where A is the amplitude of embedded sinusoidal flicker signal and theta is the orientation of the tagged information. See **Figure 2c** for an illustration of this type of power filter. Effectively, this filter p and down regulated information at oblique orientations according to the phase of the flicker signal, while leaving information cardinal angles unmodulated over time. Note that the filter was applied to the full frame, and any natural scene video will, by definition, also vary in orientation and spatial frequency power over time and space. However, as these natural variations are stochastic in nature, averaging the neural response across successive video presentations allows for the exclusive measurement of signals time-locked to the embedded flicker signal (e.g., those evoked by the embedded changes in contrast of specific orientations and spatial frequencies). After applying the filters, we reconstructed the luminance information using the modulated spatial frequency pyramid. In turn, this new “Value” channel (HS**V**) was recombined with its original hue and saturation channels and converted back to RGB colour space for presentation during the experiment. See **Figure 2f** for an example of one such modulated frame, noting that different orientations and spatial frequencies will be more relevant in different frames depending on the interactions of the two embedded frequency tags. Each raw video clip was subjected to this procedure twice, once to tag relatively higher spatial frequencies at 7 Hz and lower spatial frequencies at 9Hz, and a second time to tag relatively higher spatial frequencies at 9 Hz and lower spatial frequencies at 7Hz. See **supplementary materials** to view the spatial frequency tagged videos.

### Behavioural Visual function testing

All AMD patients and healthy controls underwent behavioural visual function testing to assess visual acuity (logMAR) and contrast sensitivity (logCS) using the Freiburg vision test (FrACT, Bach, 2024, 1996). Testing was performed from a viewing distance of 93cm.

### Display computer specifications

All displays were presented at a viewing distance of 64 cm on Alienware 27 inch AW2721D monitor with a refresh rate of 120 Hz and resolution of 2560×1440. Stimuli were presented using custom software incorporating PsychoPy video presentation software (Peirce et al., 2019). The experiment was run on a Lenovo Thinkpad laptop and Lambda workstation.

### EEG recording

EEG data were sampled at 1000 Hz using a BioSemi Active Two amplifier (BioSemi, Amsterdam, Netherlands) from 68 active Ag/AgCl scalp electrodes arranged according to the international standard 10-20 system for electrode placement in a nylon head cap^66^. Four electrooculography (EOG) electrodes were used to record eye-movement muscle artifacts. The Common Mode Sense (CMS) and Driven Right Leg (DRL) electrodes were placed to the left and to the right of POz, respectively. Eye movements were recorded using the Tobii Pro Fusion 250Hz eye-tracking system and synchronised with the EEG data using Lab Streaming Layer (LSL, Wang et al., 2023).

### EEG Analysis

*Pre-processing.* EEG data were pre-processed for offline analysis using the MNE-Python package (Gramfort et al., 2013). Noisy electrodes, identified via visual inspection, were replaced with cubic-spline interpolation based on the nearest channels. EEG data were average-referenced and bandpass filtered using a zero-phase bandpass filter from 1 - 100 Hz. Additionally, a notch filter at 60 Hz was applied to eliminate line noise. An independent component analysis (ICA) with 20 components was used to eliminate oculomotor artifacts, using the MNE-Python automated algorithms to identify and exclude components likely to be blink artifacts, eye-movement artifacts, or muscle noise.

*SSVEP analysis.* For each 7.5s video, we computed a 7s epoch beginning 0.5s after video onset. For each of these epochs, EEG data across all channels were linearly detrended and baseline corrected. Videos on which the absolute amplitude at any occipitoparietal electrode (Iz, I1, I2, Oz, O1, O2, POz, PO3, PO4, PO7, PO8, Pz, P1, P2, P3, P4) exceeded 150 *µ*V were excluded from further analysis. Using these epochs, we averaged across all videos for each of the two spatial frequency-flicker frequency conditions (lower SF 7 Hz, higher SF 9 Hz | lower SF 9 Hz, higher SF 7 Hz). Thus, we computed ERPs representing the neural activity synchronised to the embedded flickering signals, and averaged out spontaneous endogenous neural activity which changed in phase across successive video presentations. These ERPs were submitted to Fast Fourier Transforms (FFTs), and SSVEPs were taken as the average power across the two occipitoparietal electrodes that showed the highest SSVEP amplitude for each flicker frequency (7 Hz, 9 Hz). As flicker frequency and spatial frequency were fully counterbalanced, this approach optimized SSVEP amplitudes equally for the relatively lower and higher spatial frequencies. Signal to noise ratio (SNR) was calculated by dividing the power at each frequency by the average of the two neighbouring frequencies on either side. Given the signal length of 7 s, this corresponded to +/- 0.29 Hz. This was then converted to decibels as follows:

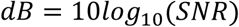

### Machine Learning

Electrode sites used in machine learning analyses were: Iz, I1, I2, Oz, O1, O2, POz, PO3, PO4, PO7, PO8, Pz, P1, P2, P3, P4. Regression was implemented using Lasso (L1) regularised regression, with parameters estimated using the union of intersects (UoI) method (Sachdeva et al., 2021). UoI Lasso regression was implemented through the PyUoI package, using the default parameters with the exception of *stability_selection* (Bouchard et al., 2017; Sachdeva et al., 2021). The *stability_selection* parameter was set to 0.5 to reduce overfitting by encouraging the selection of more electrodes as features. We used 5-fold cross validation to estimate goodness of fit on unseen data. Splitting the data into 5 groups, we predicted visual acuity for the members of each group using a model trained on data from the 4 remaining groups. Goodness of fit was evaluated on these predictions using the coefficient of determination (R^2^), confidence intervals on R^2^ were calculated as per Cohen et al., (2013). 5-fold cross validation was fit using *scikit-learn*. The K-nearest neighbours (KNN), logistic regression (LR) and multi-layer perceptron classifiers were all fit using *scikit-learn*. KNN was fit using 5 nearest neighbours. The MLP was trained with two hidden layers (sizes:10, 2) using Adam optimisation. Logistic regression was fit using L1 regularisation and the liblinear solver.

### Statistical tests

Statistical tests were conducted using the BayesFactor package in R (Package version 0.9.2+, R version 3.6.1, Rouder et al., 2012). Pairwise differences and differences between groups were assessed using the JZS *t*-test (Rouder et al., 2009). Bayes factors for main effects and interactions in Bayesian ANOVA models were assessed by comparing the full model (main effects + interaction + random effects) with the model containing all effects bar the effect of interest (Rouder et al., 2012). Bayes factors are reported with proportional error estimates unless the estimate of proportional error was less than 0.01%. Bayes factors are interpreted according to Jeffrey’s criteria (Jarosz and Wiley, 2014), as follows:

### Data and code availability

Python code used to implement data analysis and R code used to implement statistical analysis can be found at https://github.com/MIPLabCH/VENM-AMD.

## Results

### Groupwise frequency tagging results

Here, we proposed a novel method of eliciting SSVEPs by periodically altering the contrast of distinct spatial frequencies in natural scene videos. Thus, we first sought to confirm that the videos tagged using this method elicited SSVEP responses. As an initial step we investigated whether the video processing method had embedded the frequency tags as intended. To this end we submitted the processed videos to the 2D steerable wavelet decomposition, extracting the power with which different orientations and spatial frequencies were represented in each frame (**Supplementary Figure 1a**). As expected for natural scenes, we found that power peaked at the lowest spatial frequencies, with progressively higher spatial frequencies less strongly represented (1/f power distribution). Further, cardinal angles were more strongly represented than obliques (**Supplementary Figure 1b**). To interrogate the fidelity of the frequency tagging procedure we computed the average power for oblique angles at each spatial frequency in each video-frame and subjected this power to an FFT. This procedure showed that the tags were embedded as intended, with higher and lower spatial frequencies displaying peaks in the power spectra for their tagged temporal frequencies (**Supplementary Figure 1c**). As a point of interest, we performed the same procedure for cardinal angles and found that the tags were less strong, though still present (**Supplementary Figure 1c**). Thus, for future studies intending to use this procedure to investigate differences in orientation (rather than spatial frequency) representation, we recommend a sharper cutoff in the filter used to apply frequency tags across orientation than the sinusoidal taper applied here.

Once we had satisfied that spatial frequency information in the video set had been tagged as intended, we turned our attention to the EEG data. Using these data, we computed an ERP for each frequency-tagging condition (low SF 7 Hz, high SF 9 Hz | low SF 9 Hz, high SF 7 Hz), and subjected these ERPs to FFTs. Finally, we computed the SNR (dB) across the frequency spectrums output by the FFT. The grand-average frequency spectrums computed using this method revealed SSVEPs at the tagged frequencies (7 Hz, 9 Hz), for both AMD patients and controls (**Figure 3a**). We next visualised the topographic distribution of the grand-mean SSVEP response (Db) across tagged frequencies (7 Hz, 9 Hz). In line with previous frequency tagging protocols, SSVEPs were found to peak at occipitoparietal electrode sites, indicating a visual cortical response (Norcia et al., 2015; Renton et al., 2021, **Figure 3b**). Thus, we concluded that the novel method of frequency tagging was effective in eliciting SSVEP responses.

**Figure 3.**
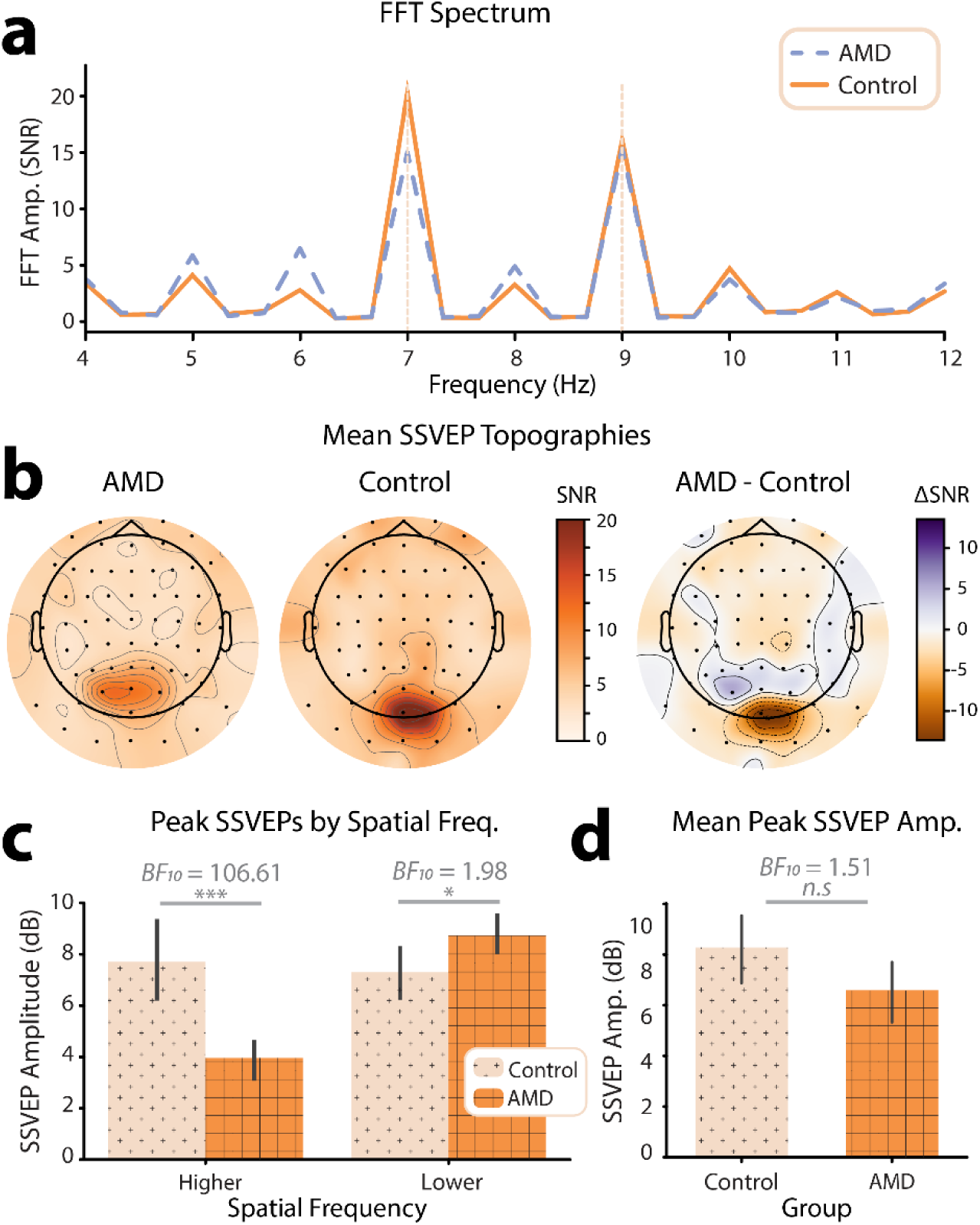
Overview of frequency tagging results. a) Grand average frequency spectrum across all participants. To generate these spectrums, ERPs to video presentations for each of the two spatial frequency/flicker frequency conditions were submitted to FFTs. For each participant, spectrums were taken as the average across the occipitoparietal electrode sites where SSVEPs peaked for each frequency. Spectrums were then averaged across all participants tagging conditions for each group (AMD, controls). Flicker frequencies (7 Hz, 9 Hz) are marked by the dashed vertical lines. b) Topographical distribution of grand-mean SSVEPs across all tagged spatial frequencies. Topographies are shown for AMD patients, healthy controls, and the difference of mean SSVEPs for AMD patients – Controls. All scalp topographies were visualized using the MNE-Python toolbox. c) Bar plot showing SSVEPs (dB) for the relatively higher and lower spatial frequencies, for AMD patients and controls. SSVEPs were taken at two occipitoparietal electrode sites for which SSVEPs peaked for each participant. d) Bar plot showing the mean SSVEPs across spatial frequencies, for AMD patients and controls. Note: Error bars on all plots indicate 95% CIs. Statistical significance is illustrated by both the Bayes Factor, and a star system representing the p value computed using frequentist statistics: n.s. p > .05, * p < .05, ** p < .01, *** p < 0.001.

Interestingly, we noted a topographical difference in the peak of the SSVEP response between AMD patients and healthy controls; on average, the SSVEP response for healthy controls peaked at the electrode sites Oz, O1, and O2. By contrast, the SSVEP response for AMD patients was shifted anteriorly, toward the electrode sites POz, PO3, and PO4 (**Figure 3b**). To investigate this further, we interrogated the topographic distribution of the SSVEP response separately for each spatial frequency (high SF, low SF). This revealed that that the topographic difference was driven by the lower spatial frequency response; while both AMD patients and healthy controls displayed a peak SSVEP response to high spatial frequencies at the electrode site Oz, the lower spatial frequency response was shifted anteriorly to POz in AMD patients. This difference in response likely reflects the retinotopic organization of the visual system; AMD patients progressively lose the ability to resolve all information in central vision, and thus this low spatial frequency information must be detected more peripherally. Indeed, such anteriorly shifted topographies have been linked to peripheral visual stimulation in a previous study aiming to retinotopically map visually evoked responses in EEG (Capilla et al., 2016). By contrast, the higher spatial frequencies tagged in these video stimuli should only be possible to resolve within the macular region of the eye and not in the periphery. Thus, in line with these findings, we would expect the neural response to high spatial frequencies to be reduced at all electrode sites in AMD patients. These topographic differences represent compelling first evidence of the efficacy of video spatial frequency tagging as a neural marker for visual field loss in AMD.

We next sought to understand how, on average, AMD patients differed from healthy controls in their SSVEP responses to the spatial-frequency tagged videos. For each participant, we computed an SSVEP amplitude for each spatial frequency by taking the average response across the two electrodes where the SSVEP response peaked for each tagged spatial frequency and flicker frequency condition (low SF 7 Hz, high SF 9 Hz | low SF 9 Hz, high SF 7 Hz). We submitted these peak SSVEP SNRs (dB) to a Bayesian ANOVA with spatial frequency (low SF, high SF) and group (AMD, control) as factors, and found decisive evidence for an interaction between these two factors (BF_10_ = 3005.81 ± 3.64%, **Figure 3c**). Note that the terminology used to describe the strength of evidence (e.g. decisive evidence) is derived Jeffrey’s criteria (Jarosz and Wiley, 2014). These criteria are outlined in **Table 1**. Interestingly, there was only anecdotal evidence for an effect of group on SSVEP SNR (dB), such that AMD patients (M = 9.61, SD = 4.29) did not differ significantly from controls (M = 11.40, SD = 3.99) in the overall magnitude of their neural response to the tagged videos (BF_10_ = 1.51 ± 23.87%, **Figure 3d**). This may be surprising, as AMD patients progressively lose the ability to resolve their central visual fields. In turn, less visual information innervates the early visual cortex, and one might expect a significantly lower amplitude neural response to the same visual information. Indeed, we found results in line with this supposition for SSVEPs to the higher spatial frequency visual information, which could only be resolved given the density of retinal ganglion cells typically found in paracentral vision. There was decisive evidence that SSVEP SNR (dB) to higher spatial frequency information in the tagged videos was weaker for AMD patients (M = 5.99, SD = 2.21) than for controls (M = 11.70, SD = 4.76, BF_10_ = 106.61, **Figure 3b**). By contrast, however, we found anecdotal evidence for the opposite effect for lower spatial frequency information. SSVEPs to lower spatial frequencies were stronger for AMD patients (M = 13.20, SD = 2.26) than for controls (M = 11.00, SD = 3.17, BF_10_ = 1.98 ± 0.01%, **Figure 3b**). This interaction between group (AMD, control) and spatial frequency response (low SF, high SF) may reflect a compensatory mechanism to the loss of central vision, such that AMD patients become more sensitive to lower spatial frequency information as they lose the ability to resolve higher spatial frequency information within the macular region.

**Table 1.**
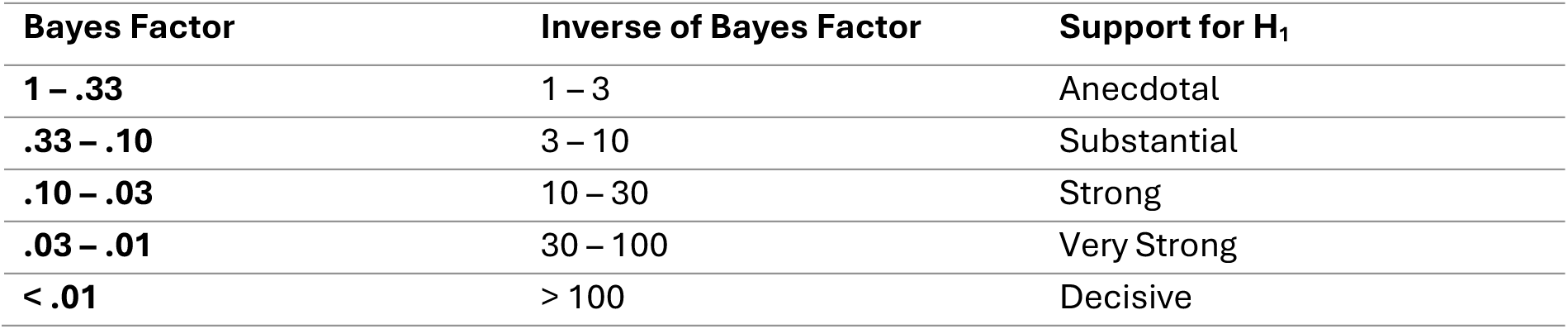
Jeffrey’s criteria for Bayes Factor inference.

| Bayes Factor | Inverse of Bayes Factor | Support for H<sub>1</sub> |
| --- | --- | --- |
| **1 – .33** | 1 – 3 | Anecdotal |
| **.33 – .10** | 3 – 10 | Substantial |
| **.10 – .03** | 10 – 30 | Strong |
| **.03 – .01** | 30 – 100 | Very Strong |
| **< .01** | > 100 | Decisive |

Given that AMD patients differed from healthy controls in their responses to both higher and lower spatial frequencies, we sought to combine these effects into a single neural marker by computing the ratio of SSVEPs to lower relative to higher spatial frequencies. Initial visual inspection, confirmed by a Shapiro-Wilk’s test for normality (W =0.78, p < .001), suggested that these ratios were heavily right skewed. We therefore computed the log of the SSVEP ratios, resulting in normally distributed data as confirmed by a non-significant Shapiro-Wilk’s test (W =0.94, p = .073). The topographical distribution of the SSVEP ratios log(low SF/high SF) suggested that this neural marker differentiates well between AMD patients and controls (**Figure 4a**): healthy controls showed a weak negative log(ratio) centred posteriorly on the scalp and peaking at electrode O2. By contrast, AMD patients showed a strong positive log(ratio) centred more anteriorly on the scalp and peaking around electrode PO4. The slight right-shifted asymmetry of these scalp topographies is a common finding in visual perceptual studies and likely reflects the underlying asymmetries in visual attention networks (Corbetta and Shulman, 2002). To confirm these observed differences, we computed the log(ratio) of the peak neural response to lower vs. higher spatial frequencies (**Figure 4b**); i.e., SSVEPs for each spatial frequency were computed at the electrode sites where they peaked (as for the results reported above). Using these ratios of lower to higher spatial frequency responses, we found decisive evidence that the log(ratio) was higher for AMD patients (M = 1.67, SD = 0.61) than controls (M = -0.14, SD = 1.23, BF_10_ = 940.47. Thus, the novel spatial frequency video tagging method allowed for the derivation of a neural marker which was sensitive to differences in visual processing between AMD patients and healthy controls.

**Figure 4.**
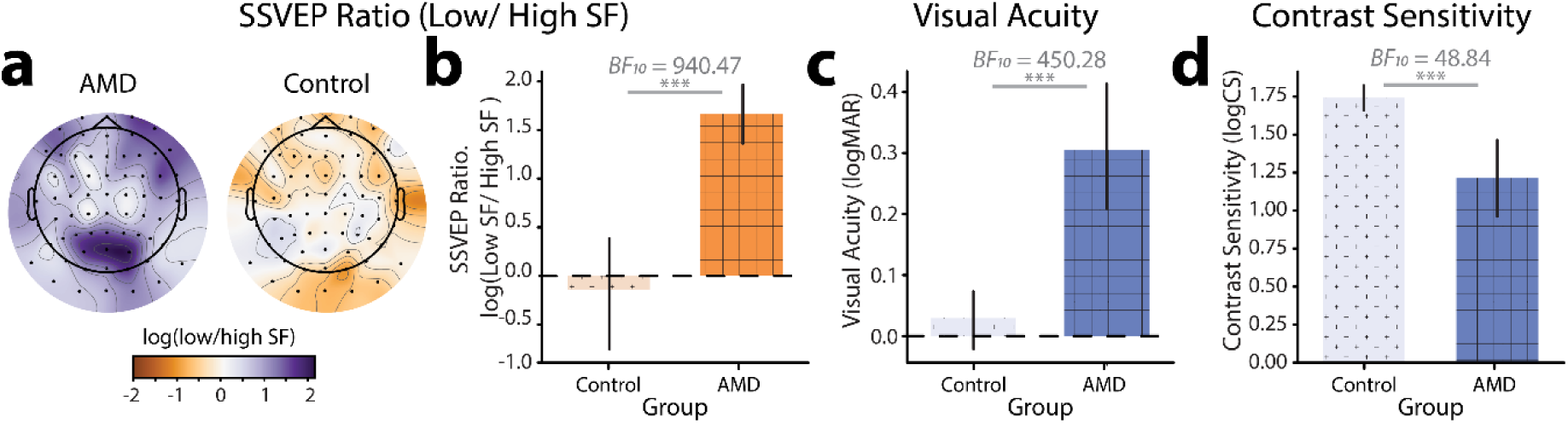
Measures of visual function. a) Topographical distribution and b) bar plot of log(Ratio) of SSVEPs to lower relative to higher spatial frequencies for AMD patients vs. controls. c) Bar plot showing visual acuity (logMAR) for AMD patients vs. controls. d) Bar plot showing contrast sensitivity (logCS) for AMD patients vs. controls.

### Individual differences: Video-evoked SSVEPs vs. Visual function

To contextualise the SSVEP results, we measured visual function in both AMD patients and controls using standardised behavioural measures of visual acuity and contrast sensitivity. The visual acuity (logMAR), contrast sensitivity (logCS), and SSVEP metrics for each participant in the study are shown in **Table 2**. As expected, we found decisive evidence that visual acuity for AMD patients (logMAR, range: 0.07– 0.81, M = 0.31, SD = 0.20) was significantly poorer than for healthy controls (range: -0.22 – 0.23, M = 0.03, SD = 0.10, | *BF_10_* = 450.28, **Figure 4c**). We also found very strong evidence that AMD patients had lower contrast sensitivity (logCS, range: 0.20 –1.94, M = 1.22, SD = 0.52) than healthy controls (range: 1.42 – 2.11, M = 1.74, SD = 2.11, | *BF_10_* = 48.84, **Figure 4d**).

**Table 2.** SSVEP and behavioural measures of visual function for each participant, sorted by visual acuity (logMAR).

|  | Group | Visual Acuity (logMAR) | Contrast Sensitivity (logCS) | SSVEP Higher SF (Db) | SSVEP Lower SF (Db) | SSVEP Ratio (log(lower/higher SF)) |
| --- | --- | --- | --- | --- | --- | --- |
| C1 | Control | -0.2122 | 1.84 | 22.08 | 7.86 | -3.27 |
| C2 | Control | -0.09 | 1.64 | 3.53 | 2.98 | -0.13 |
| C3 | Control | -0.06 | 1.94 | 10.21 | 9.07 | -0.26 |
| C4 | Control | -0.02 | 2.11 | 17.49 | 10.99 | -1.50 |
| C5 | Control | -0.02 | 1.85 | 15.33 | 8.79 | -1.51 |
| C6 | Control | 0.02 | 1.47 | 6.53 | 12.23 | 1.31 |
| C7 | Control | 0.02 | 1.80 | 13.23 | 14.46 | 0.28 |
| C8 | Control | 0.02 | 1.72 | 10.49 | 12.98 | 0.57 |
| C9 | Control | 0.05 | 1.82 | 8.71 | 11.17 | 0.56 |
| C10 | Control | 0.05 | 1.82 | 8.06 | 13.37 | 1.22 |
| C11 | Control | 0.06 | 1.57 | 13.54 | 15.11 | 0.36 |
| AMD12 | AMD | **0.07** | **1.65** | **7.95** | **10.87** | **0.67** |
| AMD13 | AMD | **0.07** | **1.94** | **6.26** | **11.52** | **1.21** |
| C14 | Control | 0.08 | 1.77 | 11.25 | 9.38 | -0.43 |
| C15 | Control | 0.09 | 1.83 | 17.87 | 14.07 | -0.88 |
| AMD16 | AMD | **0.09** | **1.81** | **6.37** | **12.89** | **1.50** |
| AMD17 | AMD | **0.11** | **1.40** | **5.16** | **13.86** | **2.00** |
| C18 | Control | 0.12 | 1.59 | 7.80 | 14.70 | 1.59 |
| C19 | Control | 0.14 | 1.69 | 11.24 | 9.80 | -0.33 |
| AMD20 | AMD | **0.20** | **1.78** | **3.56** | **11.95** | **1.93** |
| C21 | Control | 0.23 | 1.42 | 9.22 | 9.84 | 0.14 |
| AMD22 | AMD | **0.23** | **0.82** | **6.63** | **13.07** | **1.48** |
| AMD23 | AMD | **0.25** | **1.59** | **7.62** | **15.98** | **1.92** |
| AMD24 | AMD | **0.28** | **1.35** | **3.77** | **12.67** | **2.05** |
| AMD25 | AMD | **0.28** | **1.31** | **0.45** | **13.36** | **2.97** |
| AMD26 | AMD | **0.38** | **1.21** | **6.06** | **11.65** | **1.29** |
| AMD27 | AMD | **0.39** | **0.95** | **8.17** | **18.76** | **2.44** |
| AMD28 | AMD | **0.41** | **0.90** | **7.75** | **13.89** | **1.41** |
| AMD29 | AMD | **0.47** | **0.93** | **4.14** | **11.22** | **1.63** |
| AMD30 | AMD | **0.53** | **0.39** | **7.91** | **10.66** | **0.63** |
| AMD31 | AMD | **0.81** | **0.20** | **7.99** | **16.16** | **1.88** |

Having established that the log(SSVEP Ratio) of lower relative to higher spatial frequencies could be used as a neural marker of AMD, we next sought to understand how well log(SSVEP Ratio) aligned with traditional behavioural measures of visual function. Using a Bayesian regression approach, we found very strong evidence that log(SSVEP Ratio) for this tagged video set linearly predicted visual acuity across all participants (logMAR, β = 0.54, *BF_10_* = 81.03, **Figure 5a**). The metric aligned less well with contrast sensitivity (logCS), though we still found strong evidence for a linear relationship between log(SSVEP Ratio) and contrast sensitivity (logCS, β= -0.41, *BF_10_* = 7.65, **Figure 5b**). Thus, log(SSVEP Ratio) aligns well with traditional behavioural measures of visual function.

**Figure 5.**
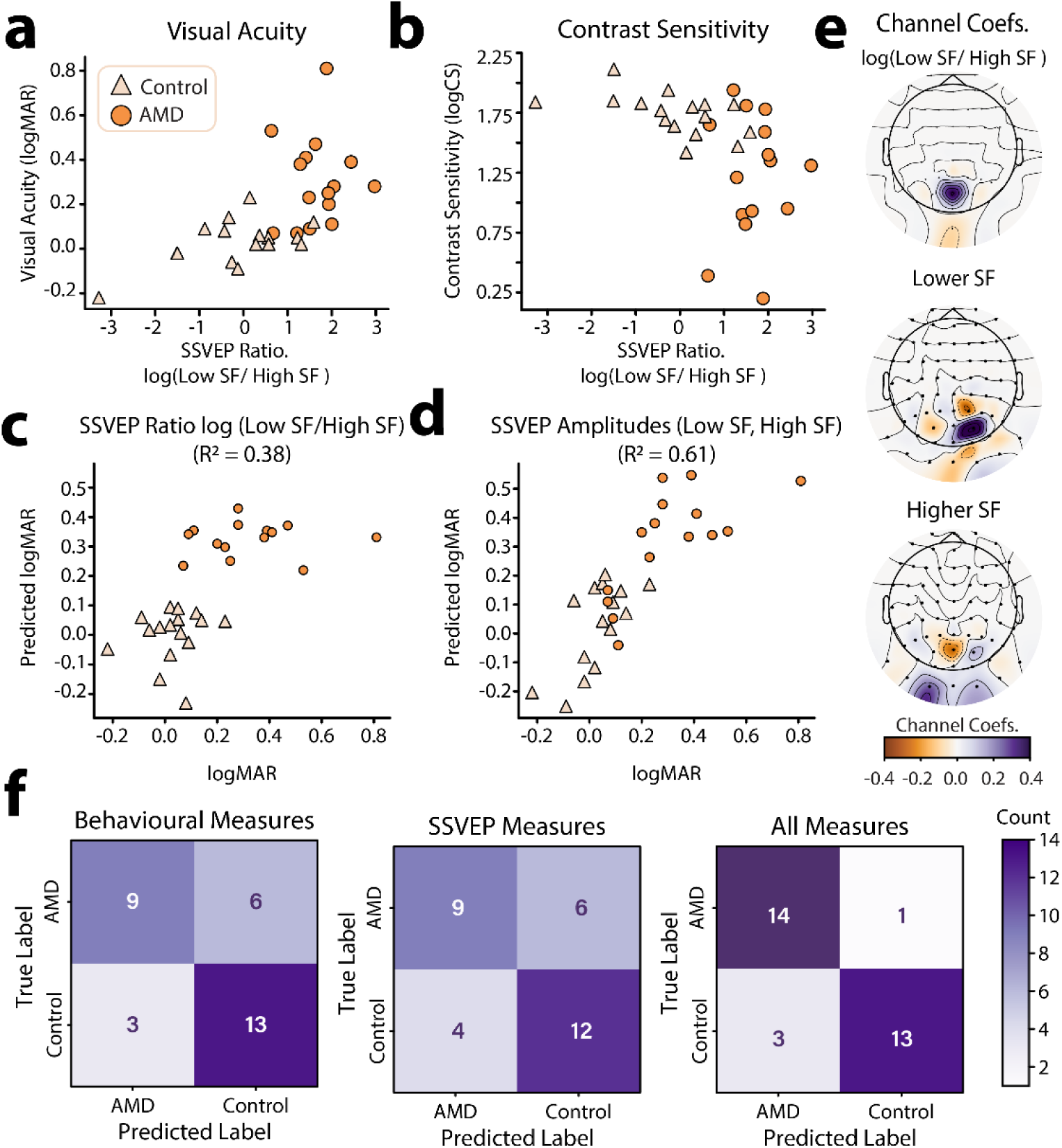
Individual differences in log(SSVEP ratio) and relationship to behavioural visual function metrics. a) log(SSVEP ratio) vs. visual acuity (logMAR) for AMD patients and healthy controls. b) log(SSVEP ratio) vs. contrast sensitivity (logCS) for AMD patients and healthy controls. c) Results of an UoI Lasso-regularised regression using group identity (AMD, Control) and the log(SSVEP Ratio) of low to high spatial frequencies at occipitoparietal electrode sites to predict visual acuity. d) Results of an UoI Lasso-regularised regression using group identity (AMD, Control) and SSVEPs to high and low spatial frequency information at occipitoparietal electrode sites to predict visual acuity (logMAR). Predicted data in c & d were acquired through 5-fold cross validation. e) EEG channel coefficients for the UoI Lasso regression shown in c & d. Topoplots were generated using “nearest” interpolation. f) Confusion matrices for the prediction of participant group (AMD, control) using behavioural visual function measures (left), SSVEP amplitudes to high and low spatial frequency information (centre), and all of the above (right) using the best performing classification model (MLP).

The log(SSVEP Ratio) metric was calculated using the SSVEPs to lower and higher spatial frequency information in the video set at the electrode sites where these SSVEPs peaked for each person. It is therefore possible that there is an underlying topographic pattern in the SSVEPs which is more strongly predictive of behavioural visual function metrics. To investigate this possibility, we used Lasso (L1) regularised regression using the union of intersections (UoI) method for parameter inference (Sachdeva et al., 2021). Lasso regularisation is known to be useful in feature selection, pushing the coefficients of less predictive features to zero. Here, we can apply this method to determine which electrode-sites were most predictive of visual acuity for higher and lower spatial frequency SSVEPs. However, application of the L1 regularisation penalty is also known to bias non-zero weights to smaller values (“shrinking “) and often incorrectly identifies non-zero weights, reducing predictive accuracy (Sachdeva et al., 2021; Tibshirani, 1996). The UoI method aims to achieve both stable feature-selection and high predictive accuracy by utilising ensemble model training and separating feature selection and estimation across two steps (Sachdeva et al., 2021). We therefore used the UoI method to fit a Lasso regression to predict visual acuity (logMAR) using group membership (AMD, Control) and SSVEP SNR (dB) to lower and higher spatial frequencies across occipitoparietal electrode sites as predictors. Visual acuity was used as it was more strongly linearly predicted by log(SSVEP Ratio) than contrast sensitivity.

To establish baseline performance, we first fit a standard linear regression with group membership (AMD, Control) as the only predictor of visual acuity. Cross validated goodness of fit for this model was R^2^ = 0.33, 95% CI [0.08, 0.57]. By contrast, a UoI-Lasso model which included the log(SSVEP Ratio) at each occipitoparietal electrode-site as predictors provided a better and more reliable fit, R^2^ = 0.38, 95% CI [0.27, 0.49] (**Figure 5c**). However, the best fitting model was the UoI-Lasso model which included individual SSVEPs to low and high spatial frequencies as predictors, providing the best and most reliable prediction of visual acuity, R^2^ = 0.61, 95% CI [0.59, 0.63] (**Figure 5d**). These results show that not just the ratio, but also the topographic distribution of SSVEPs to high and low spatial frequencies was predictive of visual function. To estimate how these topographic patterns contributed to the prediction of visual acuity scores, we computed the mean of the coefficients across the 5 cross-validation folds. These average coefficients are plotted topographically in **Figure 5e**. The topographic plots of the coefficients show that in general, weaker responses to higher spatial frequencies and stronger responses to lower spatial frequencies across occipitoparietal electrode sites were predictive of worse visual acuity (**Figure 5e**). This is evident in the coefficients for the log(SSVEP Ratio) regression with the presence of a strong positive coefficient at electrode site POz. Interestingly, there is also a weak negative coefficient at Iz, showing that at this posterior electrode site, a stronger log(SSVEP Ratio) was actually predictive of better visual acuity. Coefficients for low-spatial frequency weights were largely positive and peaked at the electrode PO2. Thus, stronger SSVEPs to low spatial frequencies at this site were predictive of worse visual acuity. This large positive peak was flanked by electrodes with weaker negative coefficients (Iz, PO3, P2), suggesting that stronger low spatial frequency responses at these flanking locations were associated with better visual acuity. By contrast, high spatial frequency SSVEPs were associated with a strong negative coefficient at the electrode site POZ, and weaker positive coefficients at PO2, I1, and I2. This is a near, though not exact, inverse of the pattern found for low-spatial frequency responses. Together, these results are indicative of the retinotopic remapping previously reported for AMD patients; suggesting AMD patients experience a shift toward prioritised processing of lower spatial-frequency information and a topographic change in sensitivity to high and low spatial frequency information (Baker et al., 2008, 2005; Dilks et al., 2014).

These findings suggest that SSVEPs elicited by the spatial frequency tagged video set are predictive of visual function in AMD. However, while the neural and behavioural measures share variance, they are not completely aligned; i.e., SSVEP measures do not capture all the variance in behavioural measures, and vice versa. This misalignment suggests that SSVEP measures will likely contain additional information which may be relevant to the classification of AMD. To asses this possibility, we trained several machine learning models to discriminate between AMD patients and controls based on 1) their visual function scores (logMAR, logCS), 2) their spatial-frequency evoked SSVEP amplitudes (low SF, high SF) at central occipitoparietal electrode sites (Oz, O1, O2, POz, PO3, PO4), and 3) all of the above. We selected three machine learning models for this task: a logistic regression with L1 regularisation as a linear model and K-nearest neighbours (KNN) and multi-layer perceptron (MLP) classifiers as non-linear models. The sensitivity, specificity, and accuracy of each of these models is shown in **Table 3**. Confusion plots for the MLP classifier, which performed best overall, are shown in **Figure 5f**. Overall, classification accuracy was better using the behavioural function scores (Mean performance: sensitivity: 83.00%, specificity = 72.33%, Accuracy = 76.33%) compared with SSVEP amplitudes (sensitivity: 71.67%, specificity = 70.67%, Accuracy = 71.00%) as predictors. However, the best result was for the combination of SSVEP amplitudes and visual acuity (sensitivity: 85.00%, specificity = 87.67%, Accuracy = 86.00%). Critically, we do not suggest that video-evoked SSVEPS should be used to diagnose AMD. Rather, these results highlight that the SSVEPs elicited by the spatial frequency tagging method provide additional information related to AMD, unique from that measured by behavioural visual function tests. This measure could therefore be gathered as part of forming a full picture of an individual patient’s visual function.

**Table 3.** Classification performance for the three classifiers, Logistic Regression (LR), K-nearest neighbours (KNN) and multi-layer perceptron (MLP). Cells are coloured on a gradient scale to reflect the value, with increasing colour saturation reflecting increasing values within the cell.

| *Predictor* | *Sensitivity* |  |  | *Specificity* |  |  | *Accuracy* |  |  |
| --- | --- | --- | --- | --- | --- | --- | --- | --- | --- |
|  | LR | KNN | MLP | LR | KNN | MLP | LR | KNN | MLP |
| **Behave** | 92% | 82% | 75% | 79% | 70% | 68% | 84% | 74% | 71% |
| **SSVEP** | 69% | 77% | 69% | 73% | 72% | 67% | 71% | 74% | 68% |
| **Both** | 87% | 86% | 82% | 88% | 82% | 93% | 87% | 84% | 87% |

### Test optimisation

In this first evaluation of the novel video spatial frequency tagging method for probing spatial frequency sensitivity in AMD patients, the test parameters were determined based on first principles and early pilot results. We therefore sought to investigate the influence of video content and experiment duration on the results, with the aim of aiding decision-making for future research using this method.

The 6 videos used in this test were selected with the aim of spanning a diverse range of colour, motion, perspective and subject matter. However, there likely exists a subset of video features that optimally elicit the SSVEP-ratio marker of visual function. While it is impossible to map this full feature-space with only 6 videos, we still sought to quantify how SSVEP metrics varied across this stimulus set. To this end, we computed the log(SSVEP Ratio) metric independently for each video in the set. It should be noted that this meaningfully increases the noise in the SSVEP metric, as per-video SSVEPs could only be calculated with a cell-size of 8 video repeats compared with the full 48 video repeats available when calculating SSVEPs for the full experiment. Interestingly, visual inspection suggested that the differences in log(SSVEP Ratio) between AMD patients and controls did vary by video ID (**Figure 6a**). To investigate this effect, log(SSVEP Ratio) data for each video were subjected to JZS t-tests to compare AMD patients vs. controls. The results, in order of significance were as follows: V3: *BF_10_* = 4.84, V2: *BF_10_* = 1.42 ±0.01%, V6: *BF_10_* = 1.19, V1: *BF_10_* = 0.43, V5: *BF_10_* = 0.37, V4: *BF_10_* = 0.36. Thus, only V3 elicited a log(SSVEP Ratio) with substantial evidence for a difference between groups (threshold: *BF_10_* > 3). Notably though, there was also no video which, when analysed alone, elicited a log(SSVEP Ratio) with substantial evidence for a null difference between groups (threshold: *BF_10_* < 0.33). A second variable of interest in assessing the efficacy of each video is the alignment with behavioural measures of visual function. As such, we computed the Pearson correlation between log(SSVEP Ratio) and each behavioural visual function metric (logMAR, logCS) for each video ID (**Figure 6b**). Again, we found variability in the strength of these relationships across videos. Similarly to the group difference results (AMD patients vs controls), the correlation analysis showed that the strongest alignment with behavioural measures of visual function was elicited by videos V3, V2, and V6, while the weakest alignment was for videos V4, V5, and V1. Thus, the semantic content and visual feature set of videos used in eliciting SSVEP metrics is likely to be an important variable in test performance.

**Figure 6.**
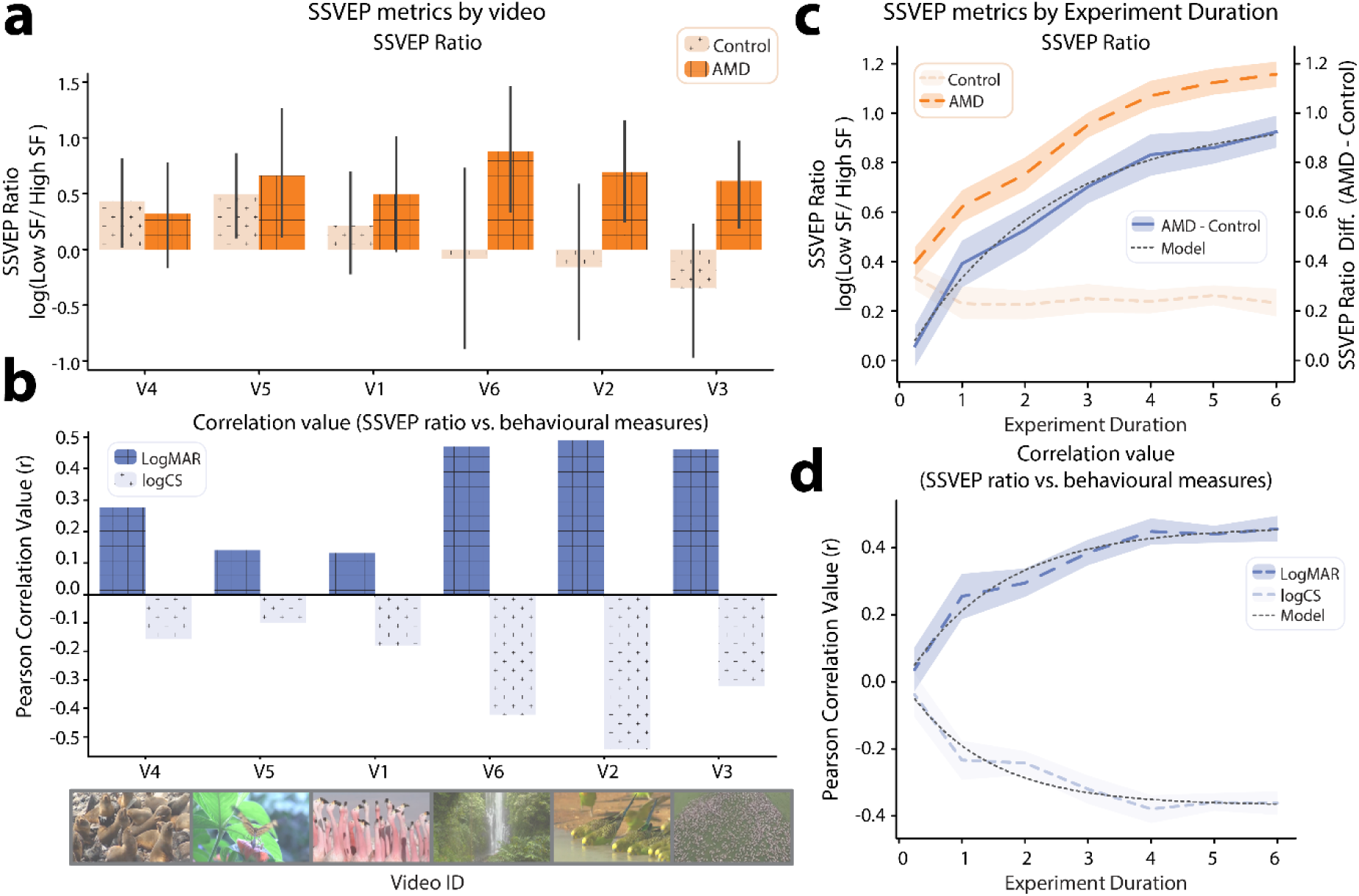
SSVEP metrics by video id and experiment duration. a) log(SSVEP ratio) between low and high spatial frequencies for AMD patients and controls, independently calculated for each video in the tagged video set. The videos are ordered on the axis from the smallest to largest mean difference between AMD patients and controls. b) Pearson correlation between log(SSVEP ratio) and behavioural measures of visual function (logMAR, logCS). The video-order is the same as in a. c) log(SSVEP ratio) between low and high spatial frequencies by simulated experiment duration. Results are shown for AMD patients, healthy controls, and the mean difference between AMD patients and controls. d) Pearson correlation between log(SSVEP ratio) and behavioural measures of visual function (logMAR, logCS) by simulated experiment duration. Shaded areas on c and d indicate 95% CI, computed by Monte-Carlo permutation.

Another important factor in future experiment design is the duration of the test. Neural responses to each probed spatial frequency range can be estimated more precisely with additional video presentation. Conflictingly, shorter diagnostic tests are valuable for both clinicians and researchers to maximise the number of distinct tests which can be performed and minimise demands on patients’ time. As such, we performed a permutation test to estimate the variance on the mean of each of the test metrics for increasing experiment durations (15 sec, 1 min, 2 min, 3 min, 4 min, 5 min, 6 min). For each experiment duration, over 30 permutations, we randomly sampled (with replacement) N/2 trials from each of the two frequency tagging conditions (low SF 7 Hz, high SF 9 Hz | low SF 9 Hz, high SF 7 Hz), where N is the number of 7.5 s trials needed to reach the targeted experiment duration. Thus, for example, in the 15 second condition, we sampled 1, 7.5 s trial from each frequency tagging condition, 30 times for each subject, to create 30 permuted samples in which each participants’ SSVEPs where computed using only 2 trials.

Using these permuted data, we calculated the mean log(SSVEP Ratio) for each group (AMD, control, **Figure 6c**). Visual inspection suggested that the log(SSVEP Ratio) for the control group converged to a stable value within 1 minute of video presentation. By contrast, the log(SSVEP Ratio) for the AMD group increased with each successive minute of video presentation. To estimate when the difference in log(SSVEP Ratio) between AMD patients and controls might converge to a stable value, we fit an inverse exponential function to this curve, as follows:

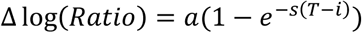

Where T is the number of training trials, a is the asymptote, s is the scaling factor and *i* is the x-axis intercept (Scolari et al., 2007). This model was found to provide a very good fit for the data (R^2^ = 0.99, **Figure 6c**). The inverse exponential model suggested that the mean log(SSVEP Ratio) difference between groups would converge to an asymptote of 0.99 after 10.5 minutes of presentation (experiment duration required to achieve 99% of asymptote value). This result suggests that a slightly longer presentation time than used here may have been optimal and should be used in future results.

Another relevant metric is that of alignment with behavioural measures of visual function. We therefore also correlated the permuted log(SSVEP Ratio) data with visual acuity (logMAR) and contrast sensitivity (logCS) data for each experiment duration. The inverse exponential model used for the log(SSVEP Ratio) difference between groups also provided a good fit for the changing r-values in this correlation as experiment duration increased (logMAR: R^2^ = 0.97, logCS: R^2^ = 0.94, **Figure 6d**). Thus, using the same model, we determined that the correlation between log(SSVEP Ratio) and visual acuity (logMAR) could be expected to converge to *r* = 0.46, within a 7-minutes experiment duration. For the correlation between log(SSVEP Ratio) and contrast sensitivity (logCS), the model suggested that an asymptote value of *r* = -0.37 could be reached within 5 minutes of experiment duration. As such, the relationship with contrast sensitivity is likely already optimally represented within the experiment duration presented here. However, effects for group differences and the relationship with visual acuity may be more strongly evident with the additional power of a longer experiment duration.

## Discussion

Here, we developed and benchmarked a neuroimaging-based visual function test which harnesses full-colour and motion videos of natural scenes. To facilitate this test, we proposed a novel technique of embedding frequency tags within video stimuli through periodic modulation of the contrast of higher and lower spatial frequencies. Neurons tuned to the tagged spatial frequencies should respond at the tagged flicker frequencies of 7 and 9 Hz, eliciting SSVEPs detectable at the scalp through EEG. Here, we demonstrated the efficacy of the novel video tagging method in eliciting SSVEP responses. In turn, we showed that these SSVEP responses were sensitive to the changes in visual processing associated with AMD and could be used as a neural marker of visual function in this patient group.

As hypothesised, we found that AMD patients differed from controls in their SSVEP amplitudes to both higher and lower spatial frequencies. The macular region of the retina is characterised by a high density of both photosensitive cone cells and the retinal ganglion cells they innervate. Retinal ganglion cells at the fovea may respond to only a single photoreceptor, as compared to thousands photoreceptors per retinal ganglion cell in peripheral vision (Watson, 2014). Thus, increasing eccentricity in the visual field corresponds with lower spatial frequency sensitivity in retina, a pattern reflected by the retinotopic organisation of spatial frequency sensitivity in the visual cortices of healthy adults. As the macula region of the retina degenerates, AMD patients progressively lose the ability to resolve higher spatial frequencies, and thus should be expected to display a decreased neural response to this information. In turn, research suggests that the visual system adapts to the loss of central vision through structural and functional reorganisation, reallocating processing resources toward lower spatial frequency information in peripheral vision (Cheung and Legge, 2005; Ramanoël et al., 2018). The SSVEP responses to the tagged video stimuli were in line with these predictions. Compared with healthy controls, AMD patients displayed a lower neural response to higher spatial frequencies across occipitoparietal electrode sites. In turn, they displayed a larger neural response to lower spatial frequencies. This lower spatial frequency response was topographically shifted toward the expected response pattern from cortical regions retinotopically mapped to peripheral regions of the visual field (Capilla et al., 2016). Thus, the SSVEPs elicited by this video set were sensitive to the functional changes in visual processing which occur in AMD.

We established that the ratio of SSVEPs to lower relative to higher spatial frequency information in the tagged video set acts as a neural marker of AMD. This marker differed significantly between AMD patients and age-matched healthy controls and could be used to predict visual acuity and contrast sensitivity in individual participants. This neural marker is likely attributable to two distinct effects in the visual cortex; the loss of afferent information from the central retina, and the neuroplastic changes in visual perceptual processing which occur in response to these lost inputs. The most profound of these adaptations is the development of a preferred retinal location (PRL), a “pseudo-fovea” in peripheral vision which is adopted when scotoma prevents central fixation. Researchers have found a large body of evidence to suggest that retinotopic remapping occurs toward the PRL. Functional magnetic resonance imaging (fMRI) data has shown that cortical regions which, in healthy adults, typically respond to visual stimulation at the fovea become responsive to visual stimulation at the PRL in long-term AMD patients (Baker et al., 2008, 2005; Dilks et al., 2014; Schumacher et al., 2008). Further, psychophysical testing has revealed changes in how visual stimuli interfere with the processing of other nearby stimuli in AMD patients (and effect termed “crowding”). Indeed, behavioural responses around the PRL begin to resemble those typically seen around the fovea (Chung, 2014, 2011). Further research has shown that visual cortical neurons whose receptive fields overlap with a central visual scotoma exhibit a shift in receptive field over time (Barton and Brewer, 2015). These receptive fields typically grow larger and shift outward from the scotoma, resulting in greater cortical sensitivity to lower spatial frequency visual information from peripheral vision as AMD progresses. Given the likely contribution these functional changes in visual processing to the log(SSVEP Ratio), it would be useful for future research to concretely map the relationship between SSVEPs, visual field loss, and time since AMD diagnosis.

The results presented here highlight the importance of neural markers of visual function. While behavioural visual function tests capture the outcome of the full process of visual perceptual processing and perceptual decision making, neural markers allow for exploration of subtle changes within specific stages of visual processing. Indeed, research suggests that the results of the behavioural visual function tests commonly administered for AMD patients often correspond poorly with AMD patients’ reported everyday visual quality of life (Broadhead et al., 2020; Taylor et al., 2016). Further, we found that AMD could be best classified using both SSVEPs and behavioural visual function scores, suggesting that these two measures explore different sources of variance in visual function. However, while these initial results are promising, the proposed method will need to be standardised and validated with a larger cohort before it can be deployed as a formal diagnostic test. Several open questions should be answered in the development of such a standardised protocol. Primarily, the feature set and semantic content of the test video set should be optimally selected. In this first study, we purposefully selected six natural scene video clips, aiming to span a diverse range of colour, motion, perspective and subject matter.

However, there likely exists a subset of video features that optimally elicit the SSVEP-ratio marker of visual function. To address these questions, we propose a neuroadaptive Bayesian optimization approach; by simulating scotoma in healthy participants, one could explore the feature space of videos which optimally elicit differences in log(SSVEP Ratio) between the scotoma vs. no-scotoma condition (Lorenz et al., 2017). Further, the optimal test duration should be established using this optimised video test set. A model trained on the current dataset suggests that 10.5 minutes should be a sufficient duration to converge on the maximal power to differentiate between AMD patients and controls using the log(SSVEP Ratio). However, this should be confirmed using a longer test duration and an optimised video test set.

Here, we presented the novel video spatial frequency tagging method in the context of diagnostic applications in AMD. However, while neural markers of visual function are useful for diagnosis and patient monitoring, they also may be useful in therapeutic applications. For example, closed-loop neurofeedback protocols aim to shift neural activity toward a pattern of activity associated with a targeted cognitive state. Advances in compute power, experimental protocols, and analytical methods over the past 15 years have meaningfully shifted the horizon of viable cognitive objectives for neurofeedback interventions. Neurofeedback protocols are rapidly developing to allow more effective and specific cognitive enhancement, with compelling evidence of training effects in visual perceptual processing; including to improve visual selective attention (Bagherzadeh et al., 2020), sustained attention (deBettencourt et al., 2015), and perceptual confidence (Cortese et al., 2020) and visual perceptual learning (Amano et al., 2016; Shibata et al., 2011). The first step in developing any such neurofeedback protocol is to identify a neural marker of the targeted cognitive objective. Neurofeedback therapy represents an especially promising treatment avenue for patients suffering from Charles Bonnet Syndrome, a common condition in AMD patients leading to vivid visual hallucinations (Singh et al., 2014; Teunisse et al., 1996). A leading hypothesis for the cause of these hallucinations is cortical hyperexcitability in the visual cortex (Bridge et al., 2024). Notably, chronic tinnitus has been attributed to the same cause in the auditory cortex (Chai et al., 2019; Langguth et al., 2007), and has been demonstrated to be treatable through neurofeedback (Gninenko et al., 2024). Previous research has shown that, due to this cortical hyperexcitability, Charles Bonnet AMD patients display profound differences in the strength of SSVEPs as compared to control AMD patients (Painter et al., 2018). Thus, SSVEPs elicited by the video spatial frequency tagging method will likely allow for a more nuanced exploration of this hyperexcitability and would provide a strong candidate for neurofeedback training aiming to regulate this maladaptive response.

Here, we presented a novel spatial frequency video tagging method to elicit neural markers of AMD, with applications in the diagnosis, monitoring, and therapeutic treatment of this patient group. This method is quick to administer, requires few electrodes, employs an ecologically valid test set of full colour and motion stimuli with natural scene statistics, does not require patients to hold fixation, and allows for exploration of subtle differences in the amplitude and retinotopy of neural responses to distinct spatial frequencies. We found that evoked SSVEPS were sensitive to differences between AMD patients and age-matched controls with normal vision, aligned well with existing behavioural measures of visual function, and represented a source of variance in visual function not computed by these behavioural tests. Finally, we explored how test parameters might be altered to optimise the power of this test in scoring visual function and recommended several avenues for future research using this method.

## Data Availability

All data produced in the present study are available upon reasonable request to the authors

https://github.com/MIPLabCH/VENM-AMD

https://osf.io/rp4q5/

## AUTHOR ACKNOWLEDGMENTS

AIR conceived of the study, AIR, JAL, and DVDV designed the study, JAL, DN, DJK, and SW coordinated the data collection, AIR analysed the data and wrote the manuscript. All authors contributed to the interpretation of results, edited the manuscript, and approved the final version of the manuscript.

## FUNDING ACKNOWLEDGMENTS

This work was funded by the American Macular Degeneration Foundation (AMDF) Breakthrough award and the Swiss Innovation Agency (Innosuisse, Project No. 116.587 IP-LS).

## COMMERCIAL RELATIONSHIP DISCLOSURE

Authors DJK, JAL, DN, AH, and SW are employees of Dandelion Science Corp. AIR and DVDV have received financial support from Dandelion Science Corp in the form of research funding. All authors are listed on a pending patent describing this work.

## Supplementary Materials

**Supplementary Figure 1.**
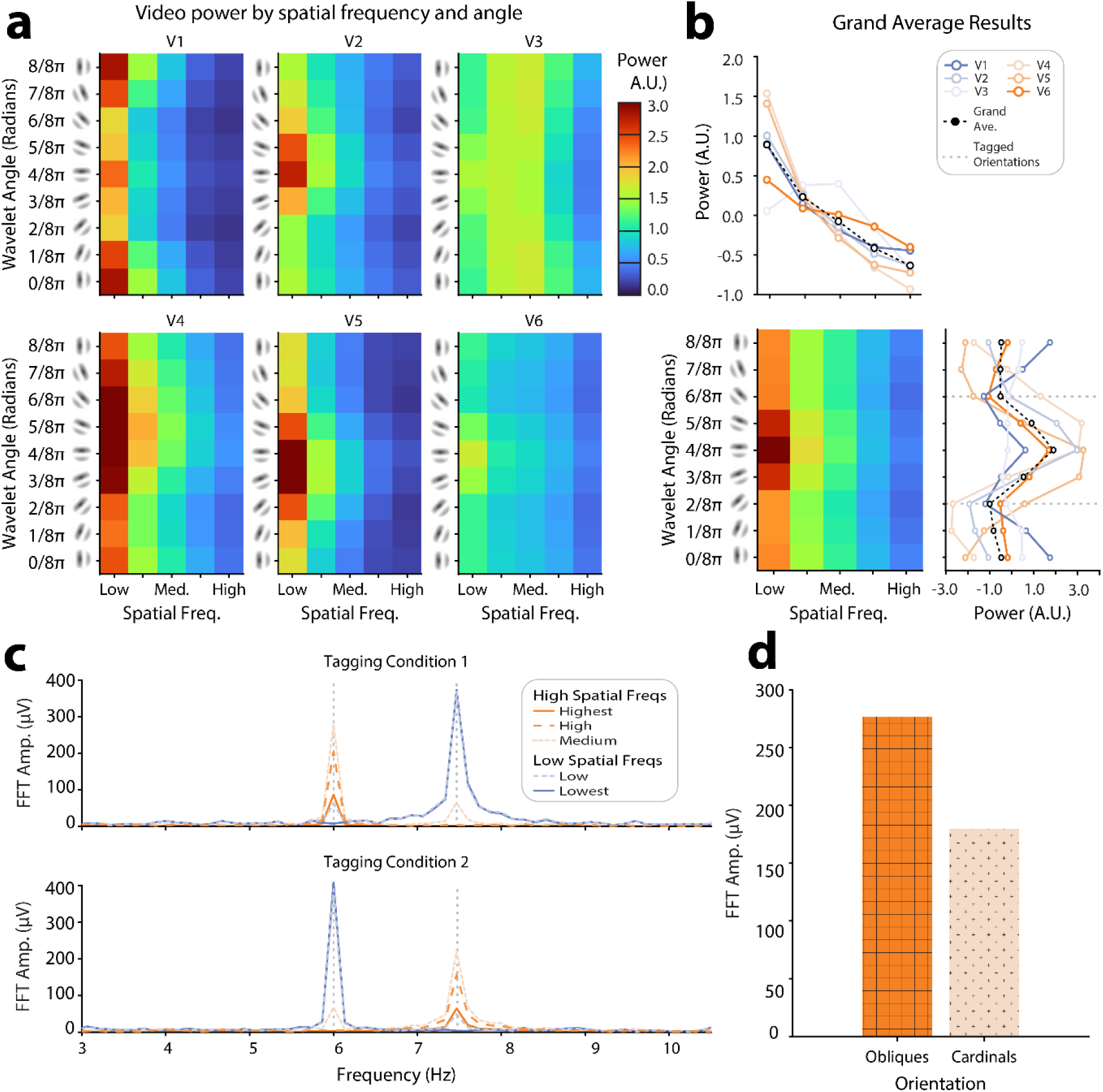
Results of the 2D Steerable wavelet decomposition of the tagged video set. a) Videos were decomposed into 5 spatial frequency bands and 8 orientation bands. The average power across all video-frames for each video in the tagged set is shown for each spatial-frequency and orientation band. b) The average power for each spatial-frequency and orientation band across all videos shows that power peaked at horizontal angles of low spatial frequencies. The axes projecting from this image plot show the video-wise variations in power across orientation and spatial frequency. Note that in these projected axes, power has been baseline corrected for each video to allow for better visualisation of the within-video variance. c) To interrogate the fidelity of the frequency tagging procedure we computed the average power for oblique angles at each spatial frequency in each video-frame and subjected this power to an FFT. The power for each spatial frequency band is shown for each of the two tagged conditions. Light-grey dashed lines show frequencies which were embedded into the videos. d) The same protocol was applied to cardinal angles. For each set of angles (oblique, cardinal), the power was taken at the tagged flicker frequency for each spatial frequency and for each tagging condition. This bar plot shows the average tagged power across these spatial frequencies and tagging conditions, for cardinal angles vs. obliques.

