## Supplementary figures and images for "Video-evoked neuromarkers of visual function in age-related macular degeneration"

### Supplementary Figure 1

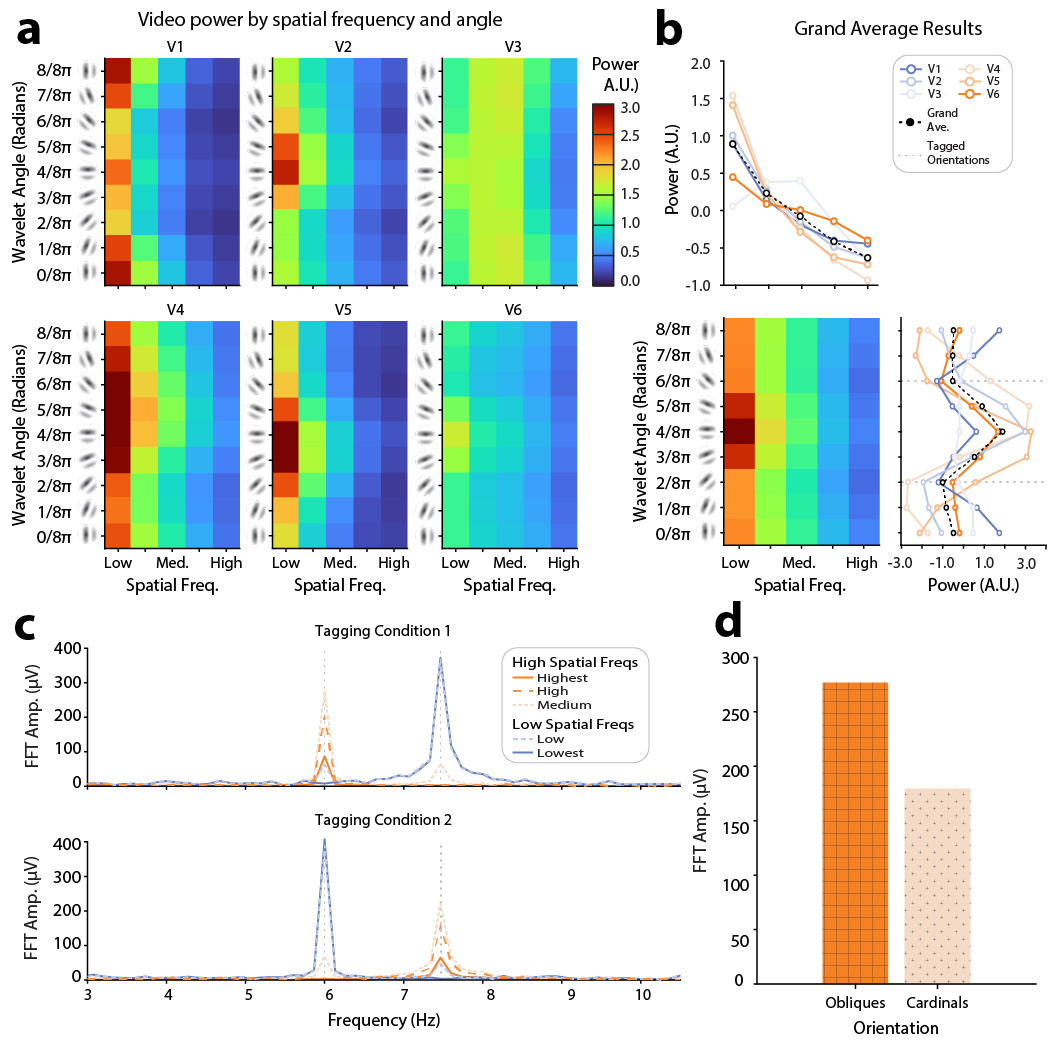
